# Comparative Thermal Effects of Single Shot Pulsed Field Ablation Systems using a Thermochromic Hydrogel

**DOI:** 10.64898/2026.06.02.26354772

**Authors:** Jaspal Singh Gill, Carlo Saija, Viral A Sagar, Zia Zuberi, Abhay Bajpai, Kawal Rhode, Lisa WM Leung, Mark M Gallagher

**Affiliations:** Department of Cardiology, Atkinson Morley Wing, St. George’s University Hospitals NHS Foundation Trust, Blackshaw Road, London, SW17 0QT UK; School of Biomedical Engineering & Imaging Sciences, Surgical & Interventional Engineering Department, St Thomas’ Hospital, King’s College London, Westminster Bridge Road, SE1 7EH

**Keywords:** Pulse-Field Ablation, Ablation Biophysics, Thermal Injury

## Abstract

**Background:** Pulse-field ablation (PFA) is regarded as a non-thermal ablation modality, but there is an increasing range of complications that could be due to thermal effects.

**Methods:** The hydrogel undergoes permanent colour change when a target temperature is reached allowing direct visualisation of the surface thermal footprint and depth. Comparative lesion sets using a variable loop circular catheter (VP), circular over-the-wire catheter (PS) and pentaspline catheter (FP) were performed. Protocols included single and stacked applications with variation of force, irrigation, and voltage. The hydrogel lesions were analysed en-face and by section using digital image analysis.

**Results:** All 3 PFA catheters tested had significant thermal footprints. The VP catheter had the largest mean surface footprint (156.1mm^2^) and thermal depth (1.31mm) compared to the other two catheters (PS 55.4mm^2^ & 1.1mm, FP 29.8mm^2^ & 1.05mm, p<0.005). Increasing irrigation showed a trend to reduce thermal footprint but did not achieve statistical significance. Increasing voltage increased thermal footprint, but increasing force had negligible effect. Stacked lesions incrementally increased thermal lesion footprint and depth in all catheters. Thermal depths of up to 2.4mm were observed. Areas of darkening and degradation of the hydrogel were observed with the VP and FP catheters, consisting of up to 47% of lesion area. No darkening was observed with the PS catheter.

**Conclusions:** There are significant thermal footprints in all the systems tested. Temperatures exceeding 60°C have been demonstrated, comparable to radiofrequency ablation, and this may explain the mechanism of injury in some reports of collateral damage during PFA.

## Introduction

Pulse-Field Ablation (PFA) is often described as non-thermal and a tissue-selective ablation modality and there has been rapid uptake in electrophysiology. Advantages of PFA include faster procedure times, and encouraging safety profiles^1,2^, whilst appearing to maintain procedural efficacy. Multiple clinical PFA systems are available, particularly as “single-shot” catheters, which facilitate rapid pulmonary vein isolation, with common examples being FaraPulse (Boston Scientific MA), PulseSelect (Medtronic MN), and Varipulse (Johnson & Johnson CA). It is important to recognize that due to each system’s proprietary PFA waveform and different catheter designs, the safety and efficacy profile of each system is unique, and results from one system therefore cannot be extrapolated to another. Individual evaluation of each system’s efficacy and safety profiles is key.

As the number of PFA procedures has increased over the past 5 years there has been a recognition of complications that would not be expected in a truly tissue-selective modality. This has included both reversible and irreversible phrenic nerve injury ^3–5^, Dressler’s syndrome/Post Cardiac Injury Syndrome^6^, and higher than anticipated rates of both overt and silent cerebrovascular events ^7–11^. The mechanisms of these complications are not fully understood. Phrenic nerve injury could possibly be due to non-tissue selective nervous system effects of PFA, and cerebrovascular events could be related to microbubble formation. Additionally, there have been reports of atrio-oesophageal fistulae using dual-energy devices, although the case specific circumstances of these have not yet been published.^12^

Some of the complications observed during PFA may be due to thermal effects. Although tissue preference of PFA waveforms can be established, significant thermal heating from PFA application will reduce tissue selectivity and shift the balance of tissue effects from irreversible electroporation to thermal necrosis. There is growing appreciation of the thermal effects in multiple different PFA systems^13^, and indeed some catheters use a temperature rise as an indication of tissue contact and successful lesion application^14^. It is possible that the thermal effects of PFA have been underestimated and the designation as “non-thermal” is inaccurate. We aim to evaluate the thermal effects of three common “single-shot” clinical PFA systems using a novel thermochromic myocardial phantom that allows direct visualization of the thermal footprint of these systems.

### Aims

1. Identify the thermal effects of PFA catheters on a hydrogel phantom model at 50°C and 60°C
2. Compare the different thermal profiles across three clinically used systems

## Methods

### Thermochromic Hydrogel

A novel thermochromic myocardial hydrogel phantom was used for testing (Peach Simulators, London, UK). The hydrogel consists of a long-chain cross-linked alginate derived hydrogel with saline hydration and impregnated with thermochromic dyes. The long chain biochemical structure of the hydrogel mimics the myofibrillar structure of myocardium. The thermochromic dyes undergo colour change when a threshold temperature is reached. The colour change is binary and is indicative of the area in which the threshold temperature is achieved, creating isothermal lines which delineate the extent of the thermal effect. Colour change is sustained, creating a “thermal footprint” and allowed detailed analysis of the size and shape of the thermal effects. Two models of hydrogel were used, one undergoing colour change at 50°C and one at 60°C. These temperature isotherms were used as at 50°C there will be clinically relevant thermal tissue damage, and at 60°C there is certainly thermal tissue effect. The hydrogels were shaped to represent a pulmonary vein antrum, allowing space for a guidewire to be advanced and allow tissue contact with the electrodes.

Thermal conductivity and electrical resistivity of the hydrogel closely matches healthy myocardium (p>0.05). OO Shore Hardness of the gel lies at the midpoint of the normal myocardial range^15^. Prior testing of radiofrequency and cryoablation lesions on the hydrogel have shown no significant difference to lesions on myocardial tissue^15^.

Therefore, both resistive and conductive heating and propagation through the hydrogel and its resulting colour change are representative of *in vivo* myocardium. These properties and previous testing of the hydrogel validate it as a model for the thermal effects of ablation.

### Apparatus and Experimental Setup

Experiments were performed in a 13 litre thermostatically controlled water bath held at 36.6°C. The large volume of fluid was used to increase the thermal mass of the setup and mimic physiological cardiovascular dynamics. Flow was maintained at 6.0L/minute representative of upper normal cardiac output.

Electrical impedance of the system was measured and maintained within the physiological range (90-130Ω) using saline. The hydrogel was held in the water bath on a rigid custom-made stand attached to the water bath.

The water bath was held on a digital scale which was zeroed between each protocol. In order to zero, the catheter was introduced into the water bath just above the hydrogel but without contact and the scale was zeroed. Subsequent force contact between the catheter and the hydrogel could then be measured to an accuracy of ±1g. The catheter was held using an applicator which facilitated transmission of force through the catheter onto the hydrogel. Force was maintained manually using the applicator tool for the duration of PFA applications. (Figure 1)

**Figure 1.**
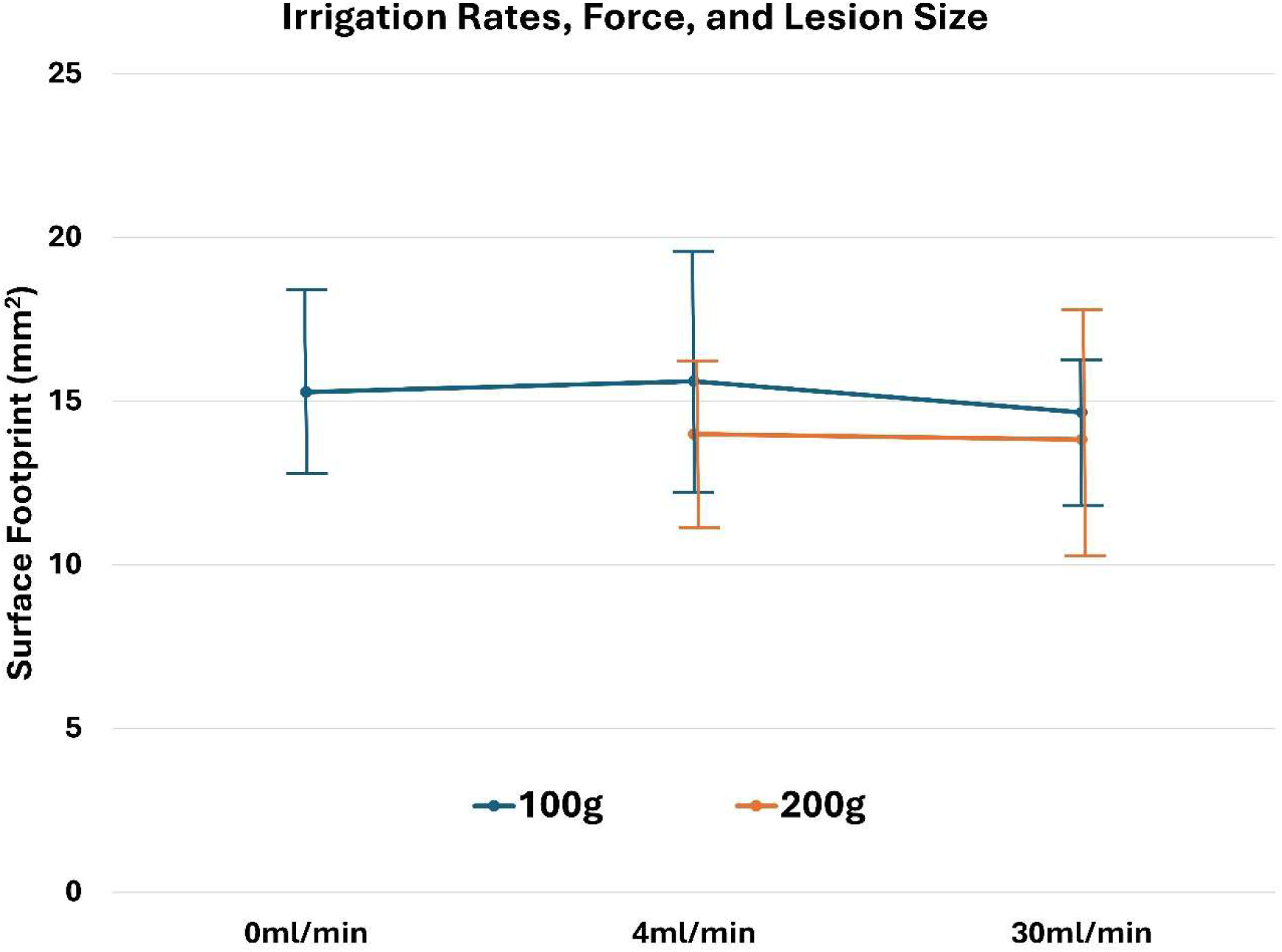
Thermal surface footprints per electrode with changes in irrigation rates and force with the Varipulse catheter.

### Catheters

Three clinically approved PFA systems were assessed. The pentaspline over-the-wire Farapulse catheter (Boston Scientific, Marlborough, MA, USA), the fixed circular over-the-wire, PulseSelect catheter (Medtronic, Minneapolis, MN, USA), and the irrigated variable loop circular Varipulse catheter (Johnson & Johnson, New Brunswick, NJ, USA). A summary of catheter characteristics is shown in Table 1.

**Table 1.**
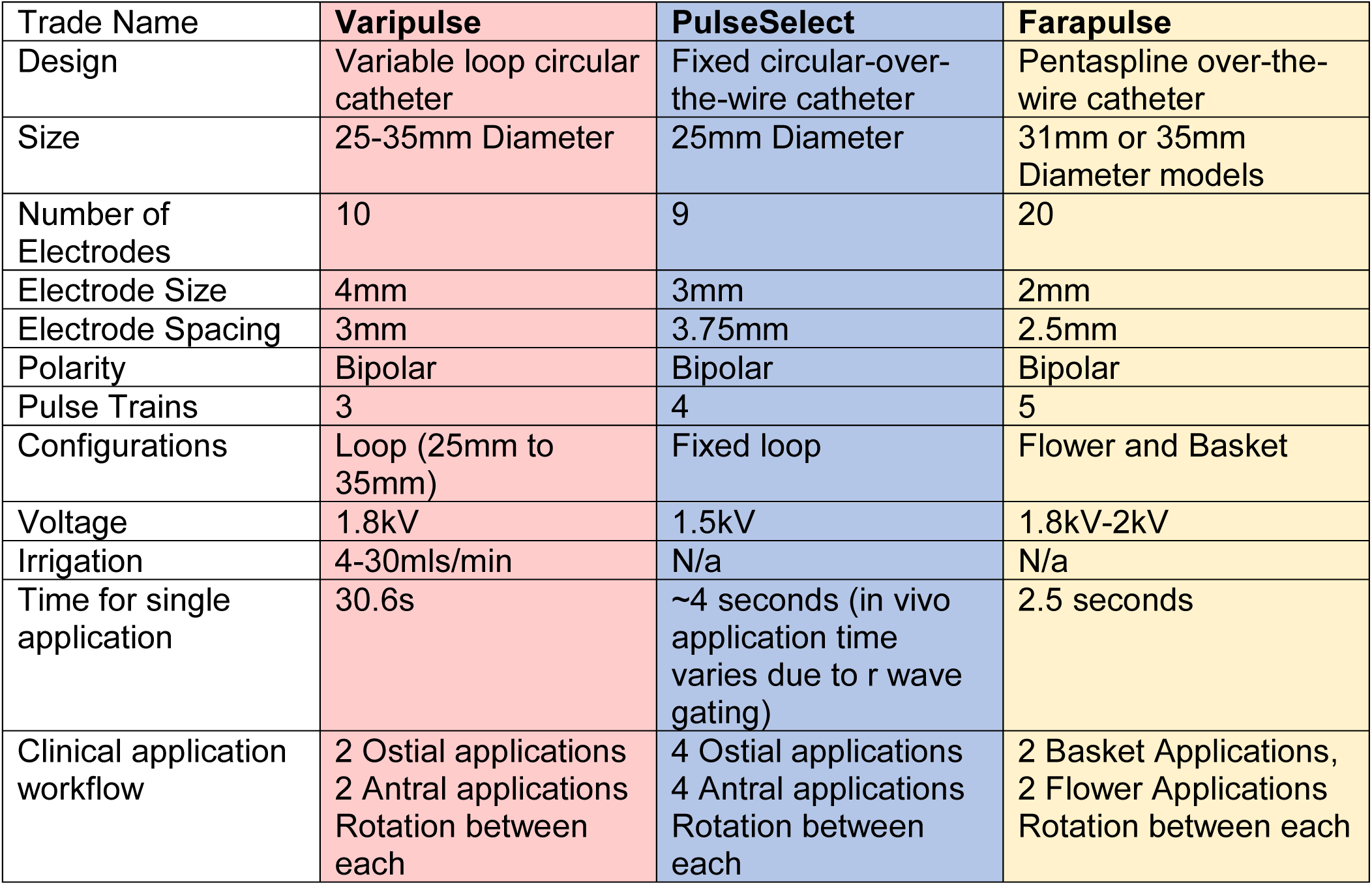
Summary table outline catheter and PFA waveform characteristics of the 3 catheters used in experimentation.

### Protocols

The thermal effects of different catheter settings and configurations were tested in the experimental protocols. The effects of force and lesion stacking were examined in all catheters. Protocols aimed to examine the settings and configurations that could be used in the clinical setting, and do not involve changes to the PFA waveforms. A full list of experimental protocols are listed in Appendix 1.

None of the catheters tested have in-built force sensing. In vivo force was estimated by comparing the shape of catheter as observed fluoroscopically to the force measured in the experimental setup when the same shape was observed. For examples, the “pinching” of the distal basket configuration of the Farapulse catheter when engaging a pulmonary vein, indicative of high contact with the vein ostium, corresponded to a force of 90g in our experimental setup. There was variation in the force applied for each of the catheters and this is indicative of the different sizes of the catheters and electrodes in each design. It is important to note that whilst these forces are significantly higher than those applied through 3.5mm tip catheters, which usually range between 10-20grams, due to the larger size and interfacing area for tissue contact, the overall pressure on tissue is similar. The use of fluoroscopic markers as the indicator for force was deliberate to mimic in vivo circumstances where there is no direct force measurement. When performing the experiments, the catheter was held in a position perpendicular to the hydrogel, to achieve a coaxial catheter electrode position. Ensuring contact between all electrodes and the hydrogel was not always possible due to the shape and angulation of the catheters. The experimental setup did not allow for tissue electrical data which is also used as a marker of tissue contact.

Direct stacking of applications was performed on the 60°C calibrated hydrogel in order to reduce the effects of thermal footprints from adjacent electrodes overlapping. Significant overlap would reduce the comparability of catheters that experienced overlap with those that did not as it would artefactually reduce the thermal surface footprint.

The Varipulse® is an irrigated catheter and irrigation rates of 0mls/min, 4mls/min and 30mls/min were used. The Farapulse catheter has two configurations of “basket” and “flower” which were both tested. Protocols with variation in voltage from 1.8kV and 2kV were also performed with the Farapulse catheter. Lesion stacking was also performed in all catheters. A comprehensive list of experimental protocols is available in Appendix 1.

### Analysis

The hydrogel samples were imaged en-face and in section. Image analysis software (ImageJ, National Institutes of Health, MA, USA) was used to analyse the size and shape of the lesions. Images were calibrated using millimetre measure in the image, placed adjacent to the hydrogels at the same distance from the camera, and imaged from above to minimise parallax error. Lesion footprint area, maximum width and length of individual lesions were measured, along with maximum depth in section.

### Statistical Methods

Continuous data is displayed with range, interquartile range and standard deviation. Data was compared with unpaired two-tailed Student T-tests. P values < 0.05 were considered significant. A Pearson correlation coefficient was used to examine the relationship between data.

## Results

Results demonstrate significant visible thermal effects on both the 50°C and the 60°C calibrated hydrogels in all 3 catheters tested. In all catheters the thermal area and depth was smaller at 60°C than 50°C.

### PulseSelect

An average thermal surface footprint was 8.2mm^2^ and thermal depth was 1.1mm on the 50°C hydrogel. On the 60°C hydrogel, the average surface footprint was 7.6 mm^2^ per electrode, total footprint of 55.4mm^2^ and thermal depth of 0.9mm.

Stacked lesions using the PulseSelect catheter resulted in incremental increases in both thermal footprint and thermal depth in a stepwise relationship (Figure 5). Force was tested at 70 and 120grams, and did not make a significant difference to the thermal footprint or depth.

### Varipulse

Manufacturer recommendations for workflow have evolved for the Varipulse catheter, with the initial irrigation rate being 4mls/min which has recently been revised to 30mls/min. A single application at 4mls/min irrigation rate had an average surface footprint of 15.6mm^2^ per electrode, total catheter footprint of 156.1mm^2^, and depth of 1.31mm. The average footprint was 14.7mm^2^ per electrode, total footprint of 146.6mm^2^, and depth 1.4mm at 30mls/min irrigation on the 50°C hydrogel. Overall irrigation rates from 0mls/s to 30mls/min, when tested at 100g of contact force, showed a trend to reducing the surface footprint (Pearson correlation coefficient r =-0.8924) but an increase in thermal depth (r=0.2226), but these did not reach statistical significance (Figure 1). A slight negative correlation for both lesion surface footprint and depth was observed at a force of 200g. Overall, force did not significantly change either the thermal footprint size or depth (100g: Footprint 156.6mm^2^, Depth 1.31mm, 200g: Footprint 140.0mm^2^ Depth 1.30mm), though contact was required for thermal effects on the hydrogel to take place.

The use of manufacturer recommended applications, with rotation between applications such that electrodes made contact with new tissue, resulted in an almost circumferential thermal footprint (Figure 2). Directly stacked lesions resulted in incremental increases in both thermal footprint and thermal depth. There was plateauing of the increase in thermal effects after the third application (Figure 5).

**Figure 2.**
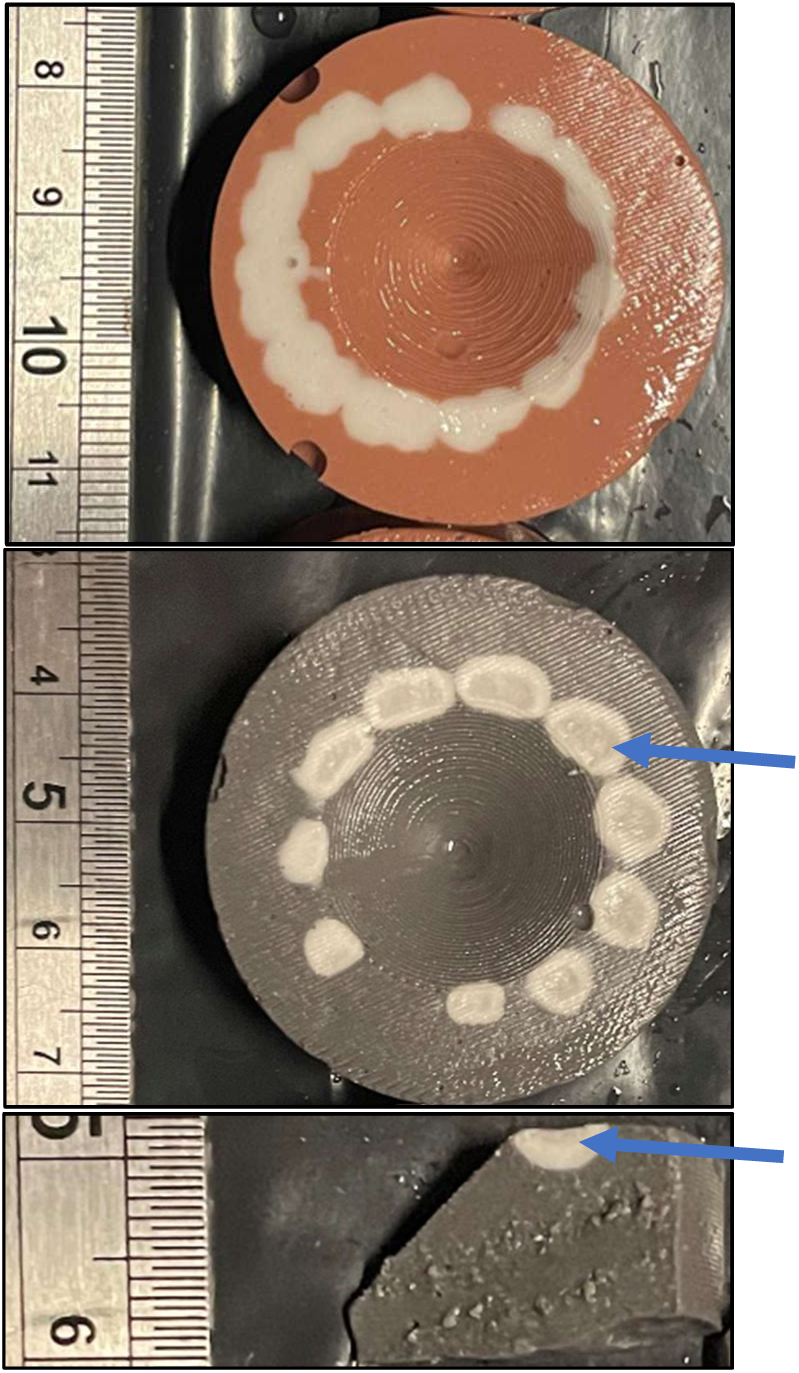
Almost-circumferential thermal footprint when a clinical workflow set of applications is delivered using the Varipulse catheter (left panel). Areas of darkening and scorch in stacked applications with the Varipulse catheter (middle panel). Section through the darkened lesions demonstrating that these changes are observed not only on the surface but also deeper in the hydrogel (right panel). The darkened areas are highlighted with blue arrows.

Darkened areas were observed immediately in the centre of the thermal footprints in stacked applications, corresponding to contact of the electrodes and subsequent areas of highest electrical field strength and highest temperature. The area of darkening consisted up to 47.0% of the area of the thermal footprint (mean 26.3%) (Figure 2). These were not observed in single applications even in the absence of irrigation and regardless of contact force, or in the clinical workflow.

### Farapulse

The Farapulse Catheter had an average thermal footprint of 29.8mm^2^ and a depth of 1.05mm^2^ when used in Flower configuration and 29.6mm^2^ and 1.08mm when used in basket configuration with the manufacturer recommended 2kV setting. Thermal footprints and depths were significantly larger at 2kV than at 1.8kV in both catheter configurations (Figure 3).

**Figure 3.**
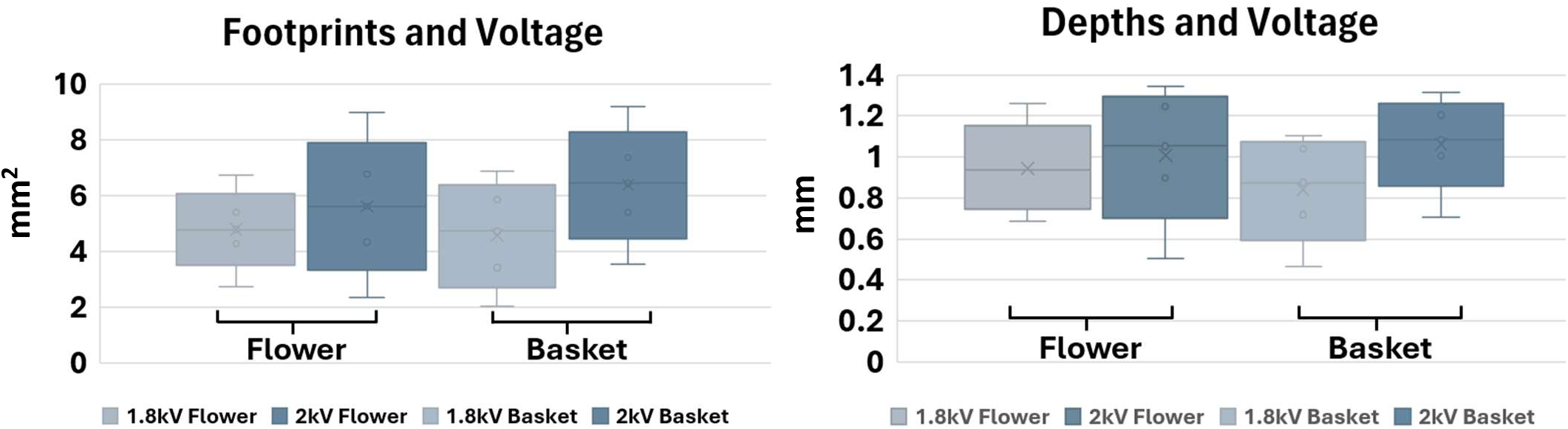
Increasing size of thermal footprints per electrode (left panel) with application stacking, and thermal depths (right panel) with stacked applications. In both Flower and Basket configuration, administration of PFA at 2kV had a greater thermal footprint than at 1.8kV.

The overall thermal footprints were slightly larger at higher forces (50g vs 100g), though this did not achieve statistical significance. In almost all the Farapulse tests, the thermal effects of only 5 electrode pairs were made, despite the catheter having 20 electrodes in total. In both basket and flower configuration, contact was only achieved with the innermost electrodes and the distal electrodes failed to make significant contact.

In a similar pattern to the previous catheters, directly stacked lesions resulted in increased thermal footprints and depths (Figure 5). Unlike in the Varipulse catheter, there did not appear to be a plateauing of thermal effects after 3 applications, and there appeared to be a more linear (Pearson correlation coefficient 0.837) relationship.

Scorching of the hydrogel was observed in stacked applications in the shape of the contact electrodes (Figure 4).

**Figure 4.**
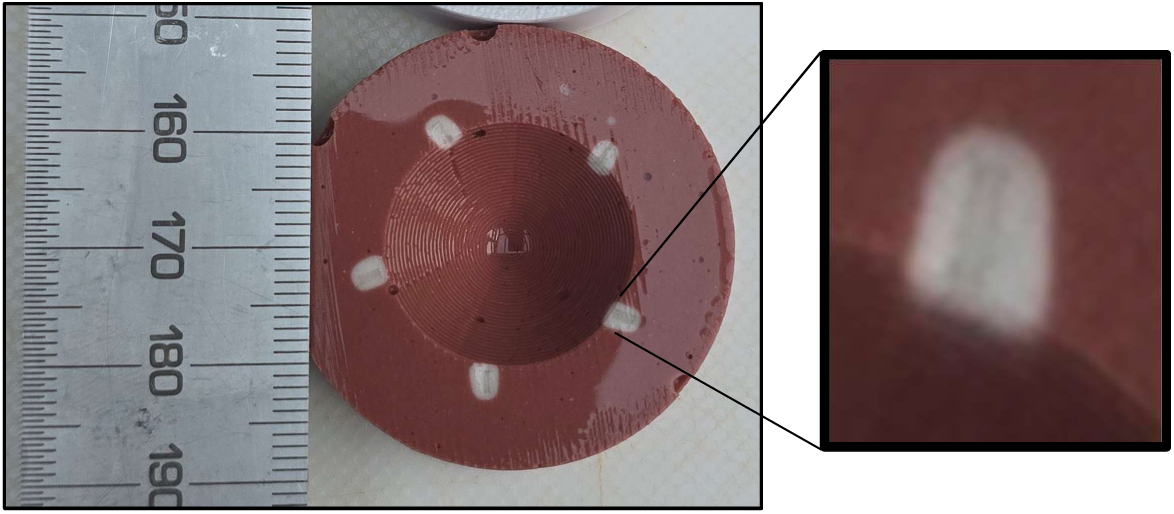
Scorching on the hydrogel as seen in stacked applications. The scorched area mimics the shape of the catheter electrode and areas of highest electrical field density.

### Comparison between catheters

There were significant differences observed in the size and depth of the thermal lesions between catheters (Figure 5). The largest thermal footprint was observed in the Varipulse catheter, and the smallest footprint was observed in the Farapulse catheter. Statistical significance with p<0.005 was observed in all comparisons.

**Figure 5.**
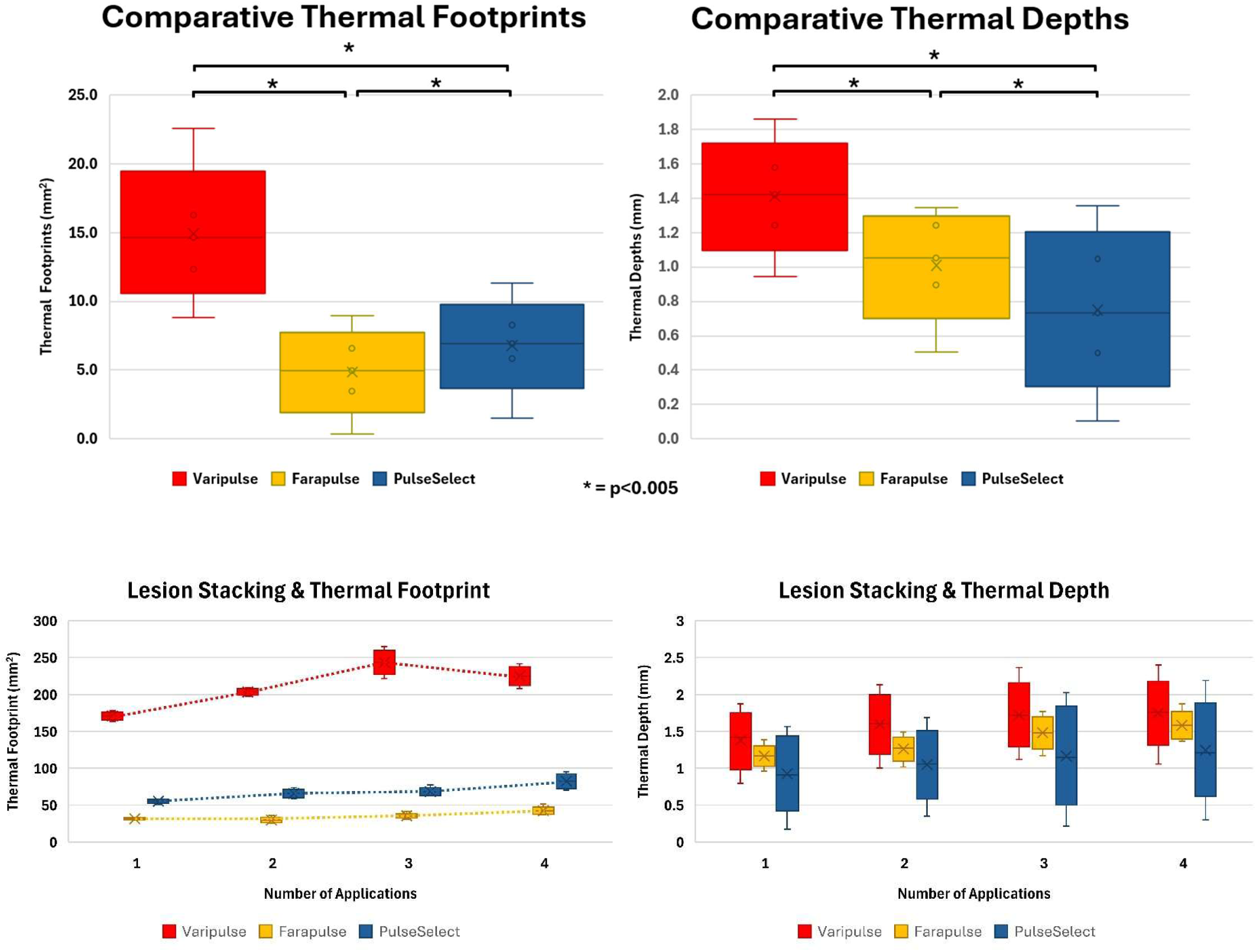
Comparative thermal footprints and depths across the three catheters tested in their clinical configurations (upper panel) and with cumulative lesion stacking (lower panel). *denotes results that achieved statistical significance with a p-value of <0.005.

The deepest thermal lesions with stacked applications were 2.4mm in the Varipulse catheter, 2.2mm in the PulseSelect catheter, and 1,8mm with the Farapulse catheter. The largest total surface footprints observed were 265.0mm^2^ with the Varipulse catheter, 95.5mm^2^ With the PulseSelect catheter and 51.8mm^2^ with the Farapulse catheter. Overall, there is over a five-fold difference in the maximum surface thermal footprint and a 33.3% difference between maximum depths between the smallest and largest catheter footprints.

Lesion stacking with the Varipulse catheter resulted in contiguous surface thermal footprints, but this was not observed in the other catheters. The shape of all thermal lesions corresponded to the shape of the contact electrodes, and in section they universally showed an ovoid morphology.

Darkened areas indicative of high temperatures and accompanying dehydration and degradation of the hydrogel and pigment were observed in the Farapulse and Varipulse catheters. In both catheters darkened areas appeared after 2 stacked applications, though the area per electrode was significantly larger in the Varipulse catheter (max 14.5mm^2^) compared to the Farapulse (max 1.38mm^2^).

## Discussion

### Thermal Effects and their Implications

These data demonstrate significant temperature rises during PFA in all systems tested. 50°C is often considered the threshold for significant thermal tissue damage, and we have observed a significant volume of the hydrogel reaching this temperature. The footprints at 60°C were also significant and the likelihood of thermal damage is more pronounced at this higher temperature. Given the validation of the hydrogel as a thermal and electrical model of myocardium, these lesions are representative of what would be observed in vivo.

Within this footprint, the balance of electroporation to thermal tissue damage is likely to be altered. Electroporation maintains extracellular structures and the profile of cell death is driven by non-inflammatory apoptosis, whereas thermal damage involves pro-inflammatory cell necrosis and destruction of tissue architecture ^16^. One of the key benefits of PFA, tissue selectivity or preference, is likely to be decreased within the thermal footprint. Normal atrial wall thickness is between 2-4mm, and given the maximum thermal depths achieved were 2.4mm, transmural thermal lesions during PFA delivery are possible. This could result in extracardiac structures being affected by thermal injury, and may contribute to cases of phrenic nerve injury observed.

The temperature rises with PFA are rapid and the results observed do not provide temporal data past the initial colour change observed. However, the size of the thermal footprint does increase for the duration of PFA delivery, coincident with the PFA pulses themselves, and it is therefore likely that high temperatures observed are for the duration of the PFA applications.

Force did not significantly change the thermal footprint in any catheter. This is consistent with the fact that force does not appear to affect the electroporated lesion size as long as there is tissue contact ^17^. This is expected given that the mechanism of PFA is dictated mostly by electrical field properties and the interaction with the tissue.

### Mechanism of Heating and Comparison with Radiofrequency Ablation

The primary mechanism of heating in PFA application is Joule heating. Joule heating describes resistive heating when current flows through a conductor. Given the short durations of PFA application, Joule heating causes a sudden increase in tissue temperature. There is minimal conductive heating due to the short duration of the PFA pulses. This contrasts to traditional radiofrequency heating where there is slower heating of the tissue over time and accompanying conductive heating to deeper tissue. In the same way that high power short duration causes shallower but wider thermal injury than traditional lower power radiofrequency ablation, PFA shifts the balance between resistive and conductive heating further creating even wider but shallower thermal footprints. (Figure 6)

**Figure 6.**
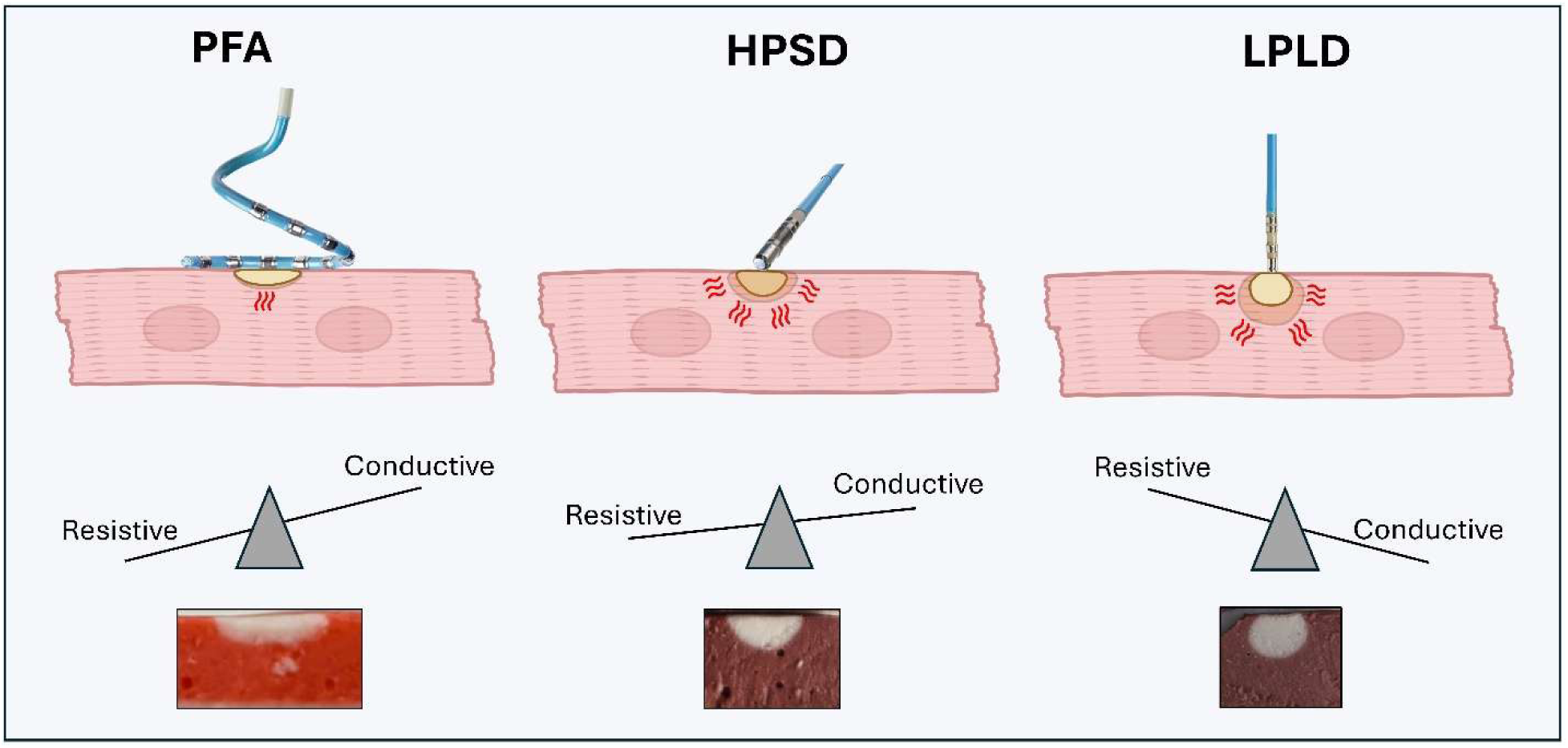
Thermal effects of PFA in terms of resistive (Joule) heating and conductive heating and comparison to the lesion shapes from previous Radiofrequency Ablation technologies. The hydrogel images shown are not to scaled comparably and are for demonstrating different lesion shapes only. LPLD = Low Power Long Duration, HPSD = High Power Short Duration.

### Comparative Effects

Comparative analysis of the catheters’ thermal footprints reveals significant differences between all. The Varipulse catheter demonstrated the largest thermal footprint and thermal depths despite irrigation. However, it is also the largest catheter, with x10 4mm electrodes. The differences between these thermal footprints are not only due to specific PFA waveforms (demonstrated by the per electrode footprint rather than the total footprint) but also due to overall catheter design elements.

The Farapulse catheter had the smallest surface footprints, despite a design of 20 electrodes arranged as 10 electrode pairs. When examining the hydrogels, we typically only observed 5 distinct thermal footprints, though a maximum of 8 (5 complete footprints and a further 3 from limited contact) were observed (Appendix 3). This was due to difficulty in achieving contact with all the electrodes with this catheter. In flower configuration, at lower forces there is contact of the proximal electrodes, but the distal electrodes fail to contact the ostium of the model. Conversely, at higher forces, there is contact of the proximal electrodes, but the high force causes deflection of the distal electrodes away from the tissue. The overall result was difficulty in achieving contact with more than 5 electrodes at any one point. Although this in part is a limitation of the model used, it is also suggestive that the Farapulse is unlikely to achieve contact of all electrodes in vivo. The basket configuration only achieves contact with the distal electrodes with this model, and this is more clearly representative of its in vivo use (Appendix 3). The Varipulse catheter however, due to the ability to bring all electrodes into contact with force application, foregoes this problem. The angulated design of the electrode loop of the PulseSelect catheter also meant that it was difficult to achieve contact of all the electrodes, and typically only 7-8 of the catheter’s 9 electrodes would have contact with the hydrogel model regardless of the force used.

Catheter design and their interaction with the model may therefore partly explain the observed results on the surface footprint. However, catheter design would be less likely to effect thermal depth observed. The maximum measured thermal depths will likely be from the catheter electrode that demonstrates the best contact, and therefore comparison of the maximum depths may allow for more inference of the thermal effect of the PFA waveform rather than the catheter design aspects.

All catheters showed an increase in thermal footprint of both size and temperature with direct stacking of applications. However, two different patterns were observed; in the Farapulse and PulseSelect catheters, the relationship appears to be more linear, with increasing the number of applications directly increasing both surface thermal footprint and the thermal depth. However, with the Varipulse catheter there is plateauing of the thermal footprint and depth after 3 applications. This is likely due to the duration of the applications and the subsequent effect that this has on thermal equilibrium reached. The Varipulse has a 30.6 second application with an obligatory “cool-off” time before a further application can be delivered, and coupled with irrigation, limits cumulative conductive heating. The tissue has more time to cool between applications and the maximum thermal footprint is therefore observed after fewer applications. Both the PulseSelect and Farapulse catheters, that have shorter applications durations of approximately 4 and 2.5 seconds respectively, see stepwise increases in thermal footprints as successive applications result in increasing surface temperatures without additional time for cooling, and subsequent conductive heating to deeper tissues.

There is the possibility that some thermal effects are indicative of effective PFA delivery, and therefore the temperature changes observed in these catheters may help predict the durability of lesions and efficacy of successful lesion placement. Further long term clinical data comparing the ablation outcomes of these catheters is needed to substantiate this possibility.

### Darkened Areas and Scorching

Experiments examining application stacking was performed with all catheters. In stacked applications (≥2) with both the Farapulse and Varipulse catheters, central areas of darkening were observed within the thermal footprints. These areas represent degradation and dehydration of the hydrogel rather than intended colour change from the thermochromic dyes. Visually, these darkened areas appeared to be scorch marks and hold a similar appearance to charring. These areas were observed on the surface with the Farapulse, but extended deeper with the Varipulse catheter (Figures 4 & 2 respectively). They were not observed with the PulseSelect catheter with any stacked applications. Although blood and the possibility of coagulum and char formation is beyond the scope of this ex vivo model, the presence of scorching on the hydrogel is suggestive that this may be an area where char could form in vivo.

The shape of the scorch marks observed followed the shape of the electrodes and electric field distribution, with scorching first observed at the edges of the electrodes, where electrical field density is highest, and the highest temperatures will be. This observation adds certainty that the darkened areas are due to excessive thermal damage.

Given the possibility of char formation and the possible consequence of cerebrovascular accident, published data on the occurrence of cerebrovascular events (CVA) in individual catheters was examined against the scorched areas. There appears to be a relationship between the darkened area and stroke rate, with increasing scorching observed with higher reported CVA rates^5,7,8,19,20^, though large registry data for the PulseSelect is lacking. The mechanism of CVA during PFA is not completely understood, though mechanisms such as microbubble formation have been proposed. These data raise the possibility of new mechanism of CVA in PFA; thermal effects giving rise to char and embolisation causing stroke.

The Varipulse catheter had initial reports of an apparent stroke rate in excess of 3%^18^, and a silent stroke rate possibly as high as 67%^7^ though after changes in workflow to reduce lesion stacking and increasing irrigation rate, this appears to have reduced significantly ^18^. This is explained by our findings, as reducing lesion stacking and increasing irrigation will reduce and possibly eliminate scorching, which would in turn be accompanied by a reduction in CVAs.

### Comparisons with other Thermal Measurements and Histological Study

Previous studies have examined thermal effects of the Varipulse catheter by using temperature probes at varying distance from the catheter and have demonstrated temperature effects of up to 58.4°C at the surface^13^. Using the hydrogel methodology, we have demonstrated temperatures in excess of 60°C, and likely significantly higher given the scorching of the tissue. An advantage of using the hydrogel to assess temperature is that it allows appreciation of isotherms, shape, and depth of thermal effects. Using temperature probe methodology also introduces inaccuracy, as the temperature probes themselves are electrical conductors, and therefore their placement within the electrical field of PFA catheter will alter the electrical field properties and the delivery of energy to the tissue. Furthermore, temperature probes are dependent on heat conduction for measurement and therefore have an inherent latency to temperature measurement.

Although early histological studies indicate that there is limited collateral damage, a more recent thorough in vitro study of immortalized cardiomyocytes exposed to electroporation supports that cellular death is in fact not due to apoptosis but due to cellular necrosis, supporting the findings of this ex vivo study that there are clinically relevant thermal effects.^21^

### Limitations

The ex vivo nature of this modelling has inherent limitations that affect its applicability in vivo. The ability to achieve contact with some catheters but not with others, and the different forces used, increases how representative the model is of in vivo PFA administration, where contact is observed fluoroscopically. However, this does limit direct mechanical comparison between the catheters. Additionally, electrical signals, another important marker of contact, were not modelled for in this setup.

The hydrogels were flat and may not completely represent the curve of left atrial vein antra. In addition, it does not model for non-myocardial cardiac tissue such as valvular apparatus, or annular tissue. Tissue microcirculation and the effects that this has on thermal effects are not accounted for in this model Fluid is recirculated at the upper limit of normal cardiac output as 6L/min. Whilst this is within normal physiological range, usual cardiac output is quoted as 5L/min. This was due to 6L/min being the lowest achievable flow rate with the experimental apparatus.

Force application was performed manually, with the intent to mimic in vivo where the catheter is held in position by the operator. However, exact maintenance of force was not possible and the force was held +-5g of the quoted force during experimentation.

As force was measured by having the entire experimental apparatus on a balance, irrigation from the catheter adds mass to the experimental setup and adds a confounder to the force being applied. Whilst this effect is negligible at lower flow rates, at higher flow rates this may introduce some inaccuracy. Additionally, the introduction of irrigant (0.9% saline) which is not the same impedance as the water bath could contribute a small inaccuracy, more significant at higher irrigation rates. However, given the large volume of the water bath (13L), the additional saline through irrigation is unlikely to significantly change to the findings, and bath impedance remained within physiological range during testing.

Blood is not modelled for in this experimental setup, and as a consequence there is no direct model for charring from the darkened and scorched areas observed. There is therefore some caution to directly link the results with char formation, however it is conceivable that these would be the areas where char would be most likely to form.

### Conclusions

These data show that commercially available pulse field ablation catheters cause a significant thermal effect during treatment applications. Thermal depths of up to 2.4mm have been observed at 60°C, and this raises the possibility of significant thermal damage rather than electroporation occurring within this footprint. There are significant differences between the thermal effects of the catheters tested and this could influence both their safety and efficacy profiles.

Char formation was known from early studies with radiofrequency ablation, however PFA’s regard as a “non-thermal” ablation modality has meant less concern with temperature effects when developing catheters. The possibility of significant thermal effects and char formation and the relationship with CVAs should be considered; workflows to reduce lesion stacking and mechanisms for temperature control in PFA catheters may help improve cerebrovascular safety. Caution should be exercised labelling PFA as a non-thermal ablation modality.

## Data Availability

Data will be shared on reasonable request to the corresponding author

## Acknowledgements

Alex Grimster, Samuel Bean, Elena Montfort Sanchez, Abigail Wiltshire, Rok Mravljak

